# Factors associated with within-individual variability of lung function for people with cystic fibrosis: a longitudinal registry study

**DOI:** 10.1101/2023.05.12.23289768

**Authors:** Marco Palma, Ruth H Keogh, Siobhán B Carr, Rhonda Szczesniak, David Taylor-Robinson, Angela M Wood, Graciela Muniz-Terrera, Jessica K Barrett

## Abstract

Lung function is a key outcome used in the evaluation of disease progression in cystic fibrosis. The variability of individual lung function measurements over time (within-individual variability) has been shown to predict subsequent lung function changes. Nevertheless, the association between within-individual lung function variability and demographic and genetic covariates is not quantified. We performed a longitudinal analysis of data from a cohort of 7099 adults with cystic fibrosis (between 18 and 49 years old) from the UK cystic fibrosis registry, containing annual review data between 1996 and 2020. A mixed-effects location-scale model is used to quantify mean FEV_1_ (forced expiratory volume in 1 second) trajectories and FEV_1_ within-individual variability as a function of sex, age at annual review, age at diagnosis, genotype and birth cohort. Mean FEV_1_ decreased with age and lung function variability showed an approximately quadratic trend by age. Males showed higher FEV_1_ mean and variability than females across the whole age range. Individuals who died during follow-up showed on average higher lung function variability than those who survived. This work opens new avenues for further research to understand the role of within-individual lung function variability in disease progression and prediction of key outcomes such as mortality.

## 1 Introduction

The availability of national registries of cystic fibrosis (CF) with longitudinal measurements of health status has led to improved understanding of disease progression and prediction of survival for the CF population. For example, longitudinal information from the UK CF registry has been used to develop dynamic prediction models of survival for people with CF [1, 2].

The evolution of lung function over time is a crucial outcome of interest in CF. In Keogh et al. [2], a measure of lung function was identified as the strongest predictor for individual survival. Although previous studies have mostly focused only on mean trajectories, some have also investigated the variability of lung function over time, giving novel insights into disease progression. For example, Morgan et al. [3] provided a working definition of within-individual lung function variability. Four-year follow-up was split into a first two-year window for evaluating lung function variability and a second two-year window for determining the association with lung function decline. This approach was used to evaluate the hypothesis that higher within-individual variability could be linked to stronger subsequent decline in lung capacity in a multicenter observational study of US individuals with CF. The authors proposed this quantification of within-individual variability instead of the count of pulmonary exacerbations to better reflect changes in lung capacity, as the latter lack a standardised definition [4]. Heltshe and Szczesniak [5] argued that such an approach does not leverage the full extent of the longitudinal information as well as the correlation between measurements taken on the same individual.

Taylor-Robinson et al. [6] instead evaluated lung function variability in the Danish CF population, splitting it into between-individual, within-individual and measurement error. Their method did not quantify variability for each individual, but identified useful thresholds to distinguish clinically relevant changes from short-range fluctuations and accounted for the long-term correlation between lung function measurement. Those findings were further explored in the US CF Foundation Patient Registry [7], to monitor disease progression at the individual level.

The aim of this work is to quantify long-term variability in lung function in the UK CF population by using the longitudinal trajectory recorded for each individual. We apply the mixed-effects location-scale model (MELSM, [8]) to specify the standard deviation as a function of covariates of interest as well as a random component which is used to account for heterogeneity that is not explained by other factors. In this sense, this study extends the previous knowledge about cystic fibrosis by quantifying the changes in within-individual lung function variability associated with age, sex and other demographic and genetic factors.

## 2 Materials and methods

### 2.1 Data source

The dataset comes from the UK Cystic Fibrosis (CF) Registry, collecting anonymised information about CF patients across the country [9]. We used the version available at the time of analysis, which includes records from 1996 until December 2020. The individuals who are clinically stable have annual reviews approximately every 12 months with the care teams in a clinic. The dataset contains data for over 13,000 individuals.

The outcome of interest in the analysis is forced expiratory volume in 1 second (FEV_1_), which counts the litres of air exhaled in the first second after maximal inspiration. This outcome was chosen instead of FEV_1_ percent predicted (the ratio between the individual FEV_1_ and the average FEV_1_ in a population of similar sex and age) to provide a more insightful interpretation of within-individual lung function variability in terms of litres rather than percentage points and to allow for comparisons at different age values.

The covariates used in this analysis are sex, age at each annual review, age at diagnosis, year of birth, homozygous F508 (or F508del) genotype. These covariates are often used in cystic fibrosis studies [2].

### 2.2 Inclusion criteria

The analysed dataset consisted of all adults between 18 and 50 years old in the CF registry with at least one recorded measurement of FEV_1_. Setting age thresholds as inclusion criteria is common in cystic fibrosis studies (e.g. [10, 11]): in particular, the older threshold is usually set to exclude cases which are less representative of the general CF population. Also, Dasenbrook et al. [10] split the analysis into childhood and adulthood by setting a threshold at 22 years old. We focused only on adults but moved the lower threshold to 18 to characterise changes in within-individual lung function variability in early adulthood.

In this work, study entry was defined as the date of the first annual review at which the age criteria were met. Individuals were followed up until death or administrative censoring at the end of December 2020. If an individual had lung transplant before or during the follow-up period, annual reviews after the transplant were excluded. Records with missing FEV_1_ and/or covariates were removed. If multiple annual reviews were recorded in a single calendar year then only data from the first one were used in the analysis.

### 2.3 Statistical analysis

The mixed-effects location-scale model (MELSM) is used to quantify within-individual variability in a longitudinal setting [8, 12]. It relaxes the standard assumption of homoscedasticity in mixed models by expressing the error variance as a function of covariates and a subject-specific random effect. The detailed formulation of the model is reported in Appendix A. The MELSM is made of three key building blocks: the mean submodel, the variability submodel and the distribution of the random effects.

The mean submodel is similar to a standard mixed model, where the outcome of interest is modelled as a function of known covariates as well as random effects (which capture the correlation of the measurements within each individual and/or each cluster of individuals). In the variability submodel, the residual standard deviation is specified as another function of known covariates as well as a subject-specific random effect. The distribution of the random effects provides a link between the two submodels. The random effects are assumed normally distributed with mean zero; their covariance measures the correlation between the random effects.

In this work, we use MELSM to evaluate changes in the mean and standard deviation of FEV_1_ as functions of sex, age at annual review, age at diagnosis, year of birth and F508 genotype. Age-sex interactions are included in both submodels, to show how the mean and variability change over time within each sex. Age at diagnosis is dichotomised into diagnosis before 1 year of age and diagnosis after 1 year. F508 genotype is also transformed into a binary covariate: 2 alleles versus 1, 0, missing or unknown. The association with age is modelled using natural cubic splines with 5 basis functions and knots located at the quantiles of the covariate values. A random intercept is included in both the mean and variability submodels, and also a correlation parameter between the mean and variability random intercepts to account for the dependence between within-individual variability and mean.

In the variability submodel, the estimated coefficients represent the association between the covariates and the variability (measured in units of log standard deviations) of the FEV_1_ measurements. To report the association on the standard deviation scale (i.e. the FEV_1_ scale), the exponential of the coefficient is computed. By combining the coefficient with the values of the covariates and the subject-specific location and scale random effects, the mean FEV_1_ and variability trajectory over time can be computed for each individual in the dataset. The average computed at each age point returns a description of the CF population mean and variability.

We fit the model in a Bayesian framework using the R package `brms` [13]. Standard prior distributions were specified (normal distribution for fixed effects, exponential distribution for random effects) with informative prior values. More details are available in Appendix A. The estimation procedure is based on a Markov chain Monte Carlo algorithm with 2000 iterations (of which 1000 warm-up). We provide the posterior estimates for the coefficients and the corresponding 95% posterior credible interval in brackets. The R code is available online at https://marcopalma3.github.io/CF_WIV/CF_WIV.html.

## 3 Results

### 3.1 Data description

The total number of individuals in the dataset analysed is 7099 with 65,522 annual reviews. The inclusion criteria are listed in Figure 3.

Table 1 describes the characteristics of the dataset by sex. Females account for less than half of the population in the sample. The distribution of individuals by number of annual reviews is displayed in Figure 4 in Appendix B. The maximum number observed is 24, which corresponds to the number of years spanned by the current version of the registry, while the mode is at 5 reviews and the median is 9 reviews. For each individual, the follow-up in years matches approximately the number of annual encounters. Figure 5 in Appendix B illustrates the changes in FEV_1_ and in the sex distribution over the age range observed in the dataset.

**Table 1:** Demographic and clinical characteristics of cystic fibrosis individuals included in the analysis. Reported are number (%) for categorical variables and median (interquartile range: 25-th percentile; 75-th percentile) for continuous variables.

|  | Females | Males | Total |
| --- | --- | --- | --- |
| Number of individuals | 3287 (46.36) | 3814 (53.64) | 7099 |
| Median number of annual reviews per individual | 8 (5; 12) | 9 (5; 13) | 9 (5; 13) |
| Median age at diagnosis in years | 0.67 (0.12; 4.04) | 0.53 (0.12; 3.97) | 0.58 (0.12; 3.99) |
| Diagnosed within the first year after birth | 1807 (54.97) | 2190 (57.42) | 3997 (56.29) |
| Homozygous for F508 | 1589 (48.34) | 1924 (50.45) | 3513 (49.47) |
| Number of individuals with one FEV<sub>1</sub> measure | 176 (5.35) | 191 (5.01) | 367 (5.17) |
| FEV<sub>1</sub> (in litres) at first annual review | 2.02 (1.41; 2.63) | 2.9 (1.98; 3.7) | 2.4 (1.64; 3.21) |
| Number of deaths | 1028 (31.27) | 987 (25.88) | 2015 (28.38) |

### 3.2 Association between covariates and lung function mean and variability

The estimates of the parameters in both mean and variability submodels of the MELSM are reported Table 2 (for categorical covariates) and Figure 1 (for continuous covariates). The FEV_1_ intercept in the mean submodel (common for all individuals) is close to 2 litres, while for the variability submodel is equal to -1.48, which corresponds to a standard deviation of *e*^−1.48^ = 0.23 litres. These represent “baseline” values before including the parameter estimates for the covariates in the model, or equivalently they refer to the case when all the covariates are set to zero.

**Table 2:** Posterior estimates with 95% CI (credible interval) for the linear fixed parameters and random effects in the model.

| Covariates | Estimate (95% CI) |
| --- | --- |
| **Mean submodel** |  |
| Intercept | 2.01 (1.99; 2.03) |
| Diagnosis after 1 year old | 0.05 (0.03; 0.07) |
| Homozygous for F508 | -0.05 (-0.06; -0.03) |
| **Variability submodel (log SD)** |  |
| Intercept | -1.48 (-1.51; -1.46) |
| Diagnosis after 1 years old | -0.04 (-0.07; -0.02) |
| Homozygous for F508 | 0.06 (0.03; 0.08) |
| **Random effect distribution** |  |
| SD for location random effects | 1.04 (1.02; 1.06) |
| SD for scale random effects | 0.45 (0.44; 0.46) |
| Correlation between random effects | 0.16 (0.13; 0.20) |

**Figure 1:**
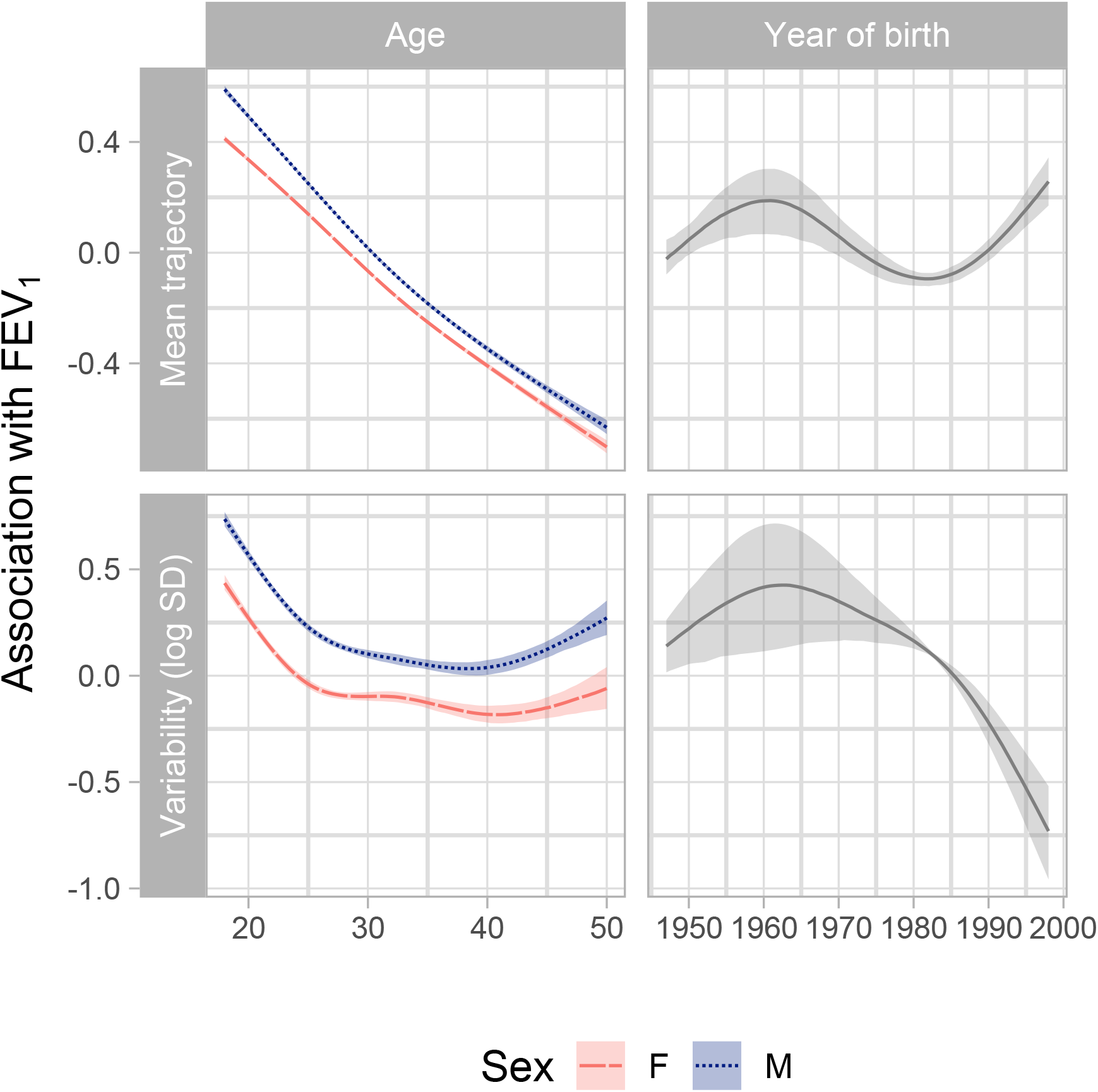
Posterior estimates of the factor-smooth interaction terms in the model described in the Equations (1) and (2) with the sex main effects added, conditional on the other covariates in the model. Top: association of age and year of birth with the mean of FEV_1_. Bottom: association of age and year of birth with the natural logarithm of the standard deviation of FEV_1_.

Those diagnosed after the first year from birth tend on average to have a higher FEV_1_ (by 0.05 litres) and lower FEV_1_ variability than those diagnosed before the first year of age after adjusting for other covariates (including the genotype). While for genotype, those homozygous for F508 tend to have lower FEV_1_ but slightly higher lung function variability than non-homozygous individuals.

Figure 1 shows the estimates of the age association for males and females. For the mean submodel, decreasing mean FEV_1_ with age is observed across the whole age range. The decrease in FEV_1_ appears to be almost linear with a slope of approximately 0.038 litres per year for males and 0.035 litres per year for females. The year of birth shows a non-linear relationship with mean FEV_1_ with the maximum reached around the cohort of 1960. Those born in this year show higher FEV_1_ on average than people born earlier or after, keeping all the other covariates constant. The minimum is obtained between 1980 and 1985: individuals in this cohort tend to have lower FEV_1_ than younger people with the same features.

In the variability submodel, for both females and males the variability tends to decrease until approximately 30 years. Between the age of 30 and 40, the variability for females show a small bounce, while for males the variability keeps declining although at a lower rate. For the cohort effect, the variability keeps increasing until the cohort between 1960 and 1965, then it decreases.

Additional heterogeneity across individuals in the lung function mean and variability is captured via the random effects in the model, as indicated by the non-zero standard deviations of the location and scale random effects in Table 2. The estimated correlation parameter indicates that those with higher FEV_1_ on average tend to show higher within-individual variability, given the other covariates in the model (as shown also in Figure 8 in Appendix B).

### 3.3 Average lung function trajectories in the CF population

For each individual in the dataset, two trajectories (one for the mean, one for the standard deviation) are fitted from the model, covering the same age interval as for the observed FEV_1_ measurements. As a simple descriptive summary of the CF adult population, the average of all the individual fitted trajectories for mean and variability are reported in solid black lines in Figure 2. The population variability for both males and females reaches the minimum at the age of 25, then it increases for older ages. In particular, for males up to 40 years old there is a stronger increase of the variability. Populations trajectories stratified by those who died during follow-up and those still alive at the end of 2020 show a higher variability for the former group across almost all the age range considered (Figure 2, bottom plot).

**Figure 2:**
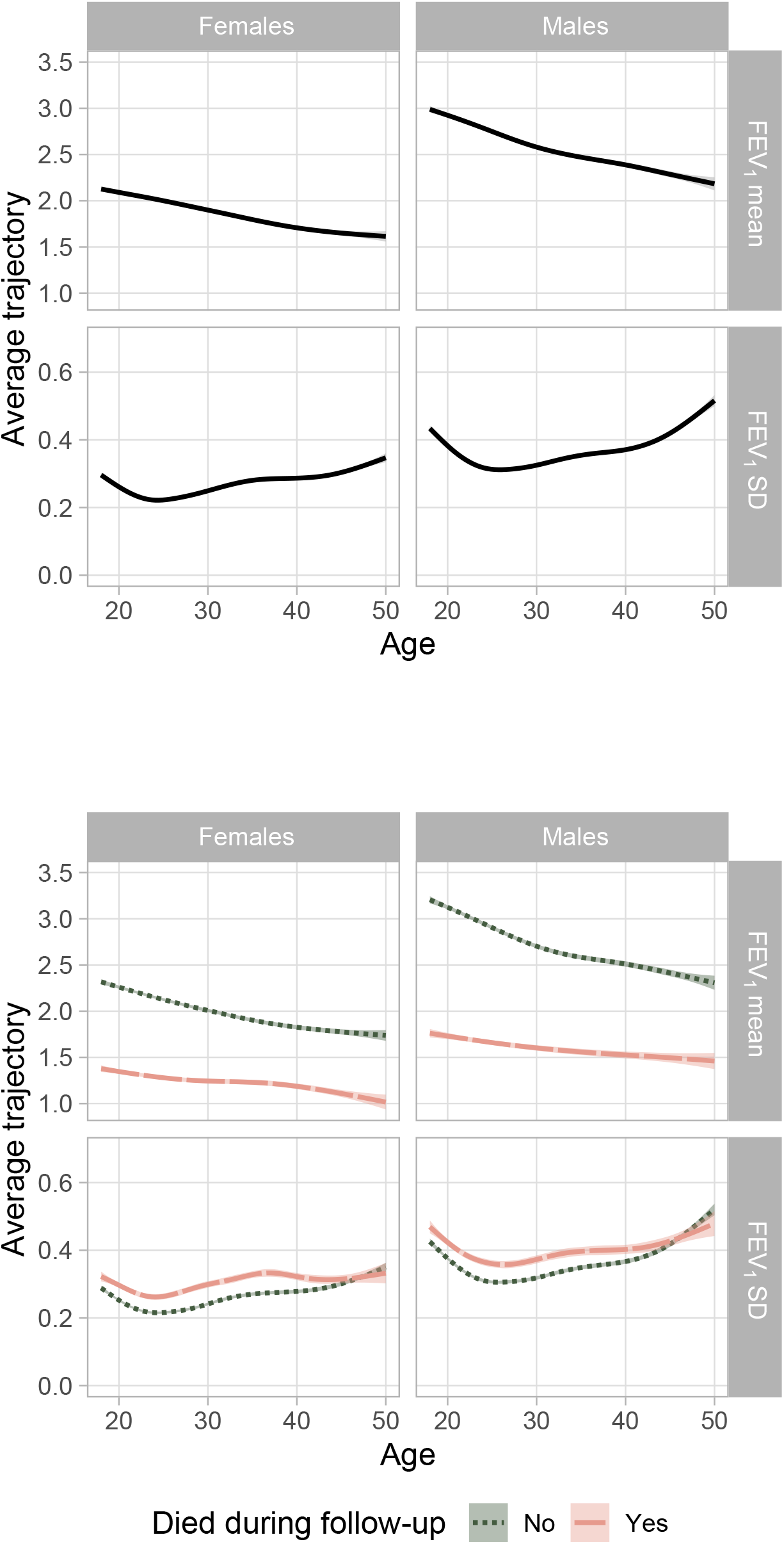
Posterior predictions of mean and standard deviation of FEV_1_ by sex and age. Top: the black solid line is the average prediction across the whole population, enclosed in the 95% prediction interval in gray. Bottom: the lines represent the average trajectory over time of variability over all individuals in the dead (orange dashed) and alive group (green dotted). The dead population includes all individuals whose death occurred before the end of 2020.

## 4 Discussion

In this work we investigated long-term (or year-to-year) lung function within-individual variability across adults in the UK CF registry dataset. The mixed-effects location-scale model allows for a flexible model of within-individual variability accounting for its relationship with covariates as well as subject-specific deviations from the population average. The results indicate that both the mean and the variability are highly heterogeneous across individuals, and only a fraction of that heterogeneity could be explained by some demographic and genetic factors.

As a first main result, we quantified the association of the covariates in the model with both mean FEV_1_ and within-individual variability. As age increases, the constant decline in mean lung function accompanies an approximately quadratic association with the variability for males, with minimal fluctuations reached around age 40. A flatter coefficient is observed for females between 25 and 33. Further work is needed to determine whether this relates to the lower life expectancy observed in the female CF population and whether it could play a role in survival prediction [14]. For both females and males, FEV_1_ variability increases after age 40, indicating higher fluctuations for older ages.

The year of birth is also associated with lung function. In older cohorts we observe increasing lung function mean as well as increasing within-individual variability, whereas younger cohorts of individuals born after 1980 show higher mean lung function and at the same time declining variability. The non-monotonic trend for this covariate suggests that there is an interplay between a left-truncation effect (as in older cohorts we observe only those healthier individual who managed to enter the study) and a cohort effect (capturing for example the improvement of treatments occurring over time), where the latter becomes the predominant driver in younger cohorts.

The second noteworthy result is that some additional heterogeneity in lung function variability is observed after accounting for the covariates, as indicated by the nonzero standard deviation of the scale random effect. This result shows that the differences between individuals are not only observed in the baseline FEV_1_, but also in the extent of the fluctuations around the mean trajectory.

Lastly, this study provides some indication that higher lung function variability could be linked to severity of diseases or more negative outcomes. For those who died during follow-up, a higher lung function variability was observed if compared to those still alive at the end of follow-up. Nevertheless, this analysis does not take into account the age at death (and the length of the individual trajectories) and therefore requires additional verification. In this direction, further study should be conducted to assess if lung function variability is a predictor of age at death, for example using joint models of longitudinal and survival outcomes accommodating within-individual variability [15]. In addition, other longitudinal outcomes in CF could be explored in the future with this framework. For example, variability in sugar levels could be added as a marker of “pre-diabetes” for individuals with CF as in [16].

We have not included infection as a covariate in the analysis, in line with previous studies of lung function variability. Our measure of variability therefore captures all components of within-individual variation, including treated and untreated exacerbations, treatment received as well as any additional variation due to e.g. CF-related diabetes or other comorbidities [3].

This study has several strengths, concerning both the specific application and the methodological aspects. To our knowledge, this is the first study to investigate factors associated with within-individual lung function variability in individuals with CF. The large cohort comprises almost all the CF population in the United Kingdom (93% of the individuals had their annual review included in the dataset in the last year [17]) and over 20 years of follow-up. A major advantage of our analysis is that the statistical approach addresses some of the limitations that emerged in the literature about within-individual variability in CF. Using the MELSM for modelling within-individual variability allows all individuals to be included in the analysis without the need to exclude those with insufficient numbers of reviews. When the number of annual reviews is small the MELSM borrows information from other individuals with similar covariate values. The definition of within-individual variability in this setting is not dependent on the definition of a baseline period and is not related to a specific summary quantity. In addition, the model flexibly accommodates nonlinear associations as for the age-sex interaction.

The same modelling approach can also be generalised to other settings. For example, by comparing the children and adult population in the UK CF dataset, one could check whether a higher lung function variability in a specific age range might be linked to different outcomes. Other extensions of the work could be tailored to CF registries from other countries, where the measurement schedule is more frequent or the number of encounters could be linked to the severity of the disease.

There are some potential limitations of the approach described in this study. The quantification of within-individual variability depends on the covariates in the mean submodel because it describes variation around the mean trajectory. Another limitation is that the model does not clearly disentangle the measurement error from clinically relevant within-individual variability, nor returns a criterion to decide whether a single observed value is “outside” some “normal” interval for an individual. Tackling these issues, along with reducing the computational time, would be useful towards a potential clinical use of within-individual variability.

The findings of this analysis suggest that modelling the within-individual variability could lead to a better characterisation of individual lung function decline in cystic fibrosis patients and potentially to improved prediction of disease outcomes.

## Data Availability

All data are available following application to the UK Cystic Fibrosis Registry Research Committee.

## Author contributions (CRediT)

Conceptualisation: all authors (equal contribution). Formal Analysis: MP. Funding acquisition: JKB, GM-T (lead contribution), RHK, AMW (supporting contribution). Methodology: MP, JKB, GM-T, RHK, AMW (equal contribution). Software: MP. Supervision: JKB. Visualization: MP. Writing – original draft: MP (lead contribution), JKB (supporting contribution). Writing – review editing: all authors (equal contribution).

## Data availability

This work used anonymized data from the UK Cystic Fibrosis Registry, which has research ethics approval (Research Ethics Committee reference number 07/Q0104/2). Use of the data was approved by the Registry Research Committee (data request 426). Data are available following application to the Registry Research Committee (www.cysticfibrosis.org.uk/the-work-we-do/uk-cf-registry/apply-for-data-from-the-uk-cf-registry).

## Funding

MP was supported by the MRC grant “Looking beyond the mean: what within-person variability can tell us about dementia, cardiovascular disease and cystic fibrosis” (MR/V020595/1). JKB was supported by MRC Unit Programme MC_UU_00002/5. RHK was funded by UK Research and Innovation (Future Leaders Fellowship MR/S017968/1). RS was supported by the National Heart, Lung and Blood Institute of the National Institutes of Health (R01 HL141286) and Cystic Fibrosis Foundation (Naren19R0 and SZCZES22AB0). GM-T acknowledges the support of the Osteopathic Heritage Foundation through funding for the Osteopathic Heritage Foundation Ralph S. Licklider, D.O., Research Endowment in the Heritage College of Osteopathic Medicine.

For the purpose of open access, the authors have applied a Creative Commons Attribution (CC BY) licence to any Author Accepted Manuscript version arising from this submission.

## Acknowledgements

We thank the Cystic Fibrosis Epidemiological Network (CF-EpiNet) Strategic Research Centre data group and Elaine Gunn for contributions to data preparation and cleaning.

## Conflict of interest

JKB has received research funding for unrelated work from F. Hoffmann-La Roche Ltd.

## Notes

### Author Declarations

This work used anonymized data from the UK Cystic Fibrosis Registry, which has research ethics approval (Research Ethics Committee reference number 07/Q0104/2). Use of the data was approved by the Registry Research Committee (data request 426).

