## Supplementary material for "Factors associated with within-individual variability of lung function for people with cystic fibrosis: a longitudinal registry study"

### A Statistical model

The final specification of the model is as follows. The mean submodel is

$$Y_{ij} = \beta_0^{\mu} + \beta_1^{\mu}\text{F508Homozygous}_i + \beta_2^{\mu}\text{AgeAtDiagnosis}_i + f_1^{\mu}(\text{Age}_{ij}, \text{Sex}_i) + f_2^{\mu}(\text{YearofBirth}_i) + \upsilon_i + \varepsilon_{ij}$$
$$i = 1, \ldots N; \quad j = 1, \ldots, n_i \quad (1)$$

The variability submodel is as follows($\varepsilon_{ij} \sim N(0, \sigma^2_{\varepsilon_{ij}})$):

$$\sigma_{\varepsilon_{ij}} = \exp\left(\beta_0^{\sigma} + \beta_1^{\sigma}\text{F508Homozygous}_i + \beta_2^{\sigma}\text{AgeAtDiagnosis}_i + f_1^{\sigma}(\text{Age}_{ij}, \text{Sex}_i) + f_2^{\sigma}(\text{YearofBirth}_i) + \omega_i\right) \quad (2)$$

Distribution of the random effects:

$$\begin{pmatrix} \upsilon_i \\ \omega_i \end{pmatrix} \sim N\left[\begin{pmatrix} 0 \\ 0 \end{pmatrix}, \begin{bmatrix} \sigma^2_{\upsilon} & \rho\sigma_{\upsilon}\sigma_{\omega} \\ \rho\sigma_{\upsilon}\sigma_{\omega} & \sigma^2_{\omega} \end{bmatrix}\right]. \quad (3)$$

The standard mixed model with random intercept is a particular case of MELSM when $\sigma^2_{\varepsilon_{ij}}$ is constant for all individuals at all measurements. In that case, $\upsilon_i$ is the only random effect in the model, accounting for the individual departure from the population average $\beta_0^{\mu}$.

The use of random effects makes the interpretation of MELSM not dissimilar from the one in standard mixed models. In the mean submodel, the estimated coefficients will quantify the association between the covariates and the lung function, keeping constant the other variables. The random effect in the mean submodel will inform about additional shifts to the individual's $\text{FEV}_1$ with respect to other individuals with the same covariate values. For example, if two hypothetical individuals share the same covariate values but the location random effect for one is 5 units higher than the other individual, then the mean $\text{FEV}_1$ for one will be 5 unit higher than the other.

Similarly, the exponential of the predicted scale random effect captures a subject-specific scaling factor (in terms of "inflation/deflation") of the $\text{FEV}_1$ standard deviation in a population with the same covariate values. For example, if the same two hypothetical individuals share the same covariate values but the scale random effect for one is 0.7 higher than the other, then the $\text{FEV}_1$ standard deviation for one would be $e^{0.7} = 2$ times the standard deviation of the other. The predicted scale random effect value provides the individual component (in addition to the common component determined by the covariate values) of the within-individual variability.

Prior predictive checks [18] induced a choice of generic informative priors:

- normal prior for linear fixed effects (all centered at 0 except the one for the intercept of the mean submodel, centered at 2)
- Exp(10) for variances of random effects and smoothing terms (which are specified in the model as random effects)
- LKJ(2) prior for random effect correlation matrix R.

The specification of the age-sex interaction in the package `brms` [13] is based on the package `mgcv` [19]. The factor-smooth interaction in `mgcv` specifies a different smoothing function for each sex with a centering constraint with respect to the model intercept. Therefore, to account for sex mean differences, a main parametric term for sex is also included. More details are available at https://stat.ethz.ch/R-manual/R-devel/library/mgcv/html/gam.models.html.

### B Additional figures

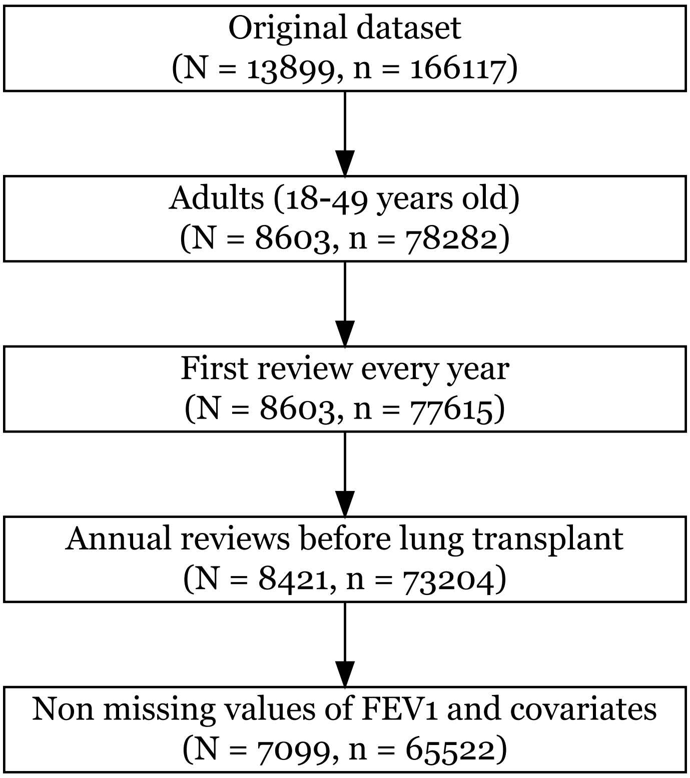


Figure 3: Diagram of inclusion criteria with number of individuals $N$ and number of observations $n$.

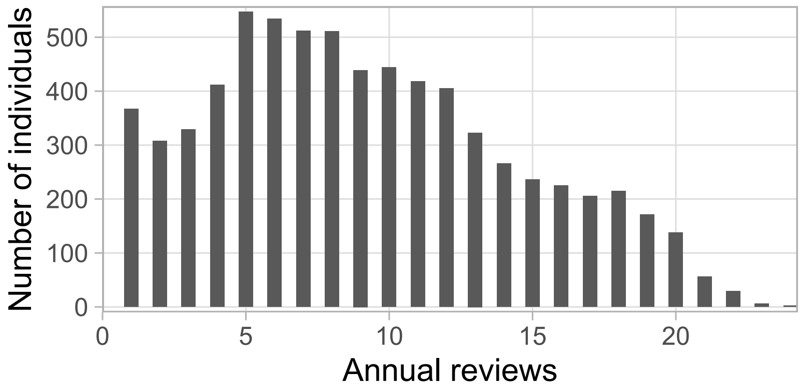


Figure 4: Barchart of individuals by number of annual reviews.

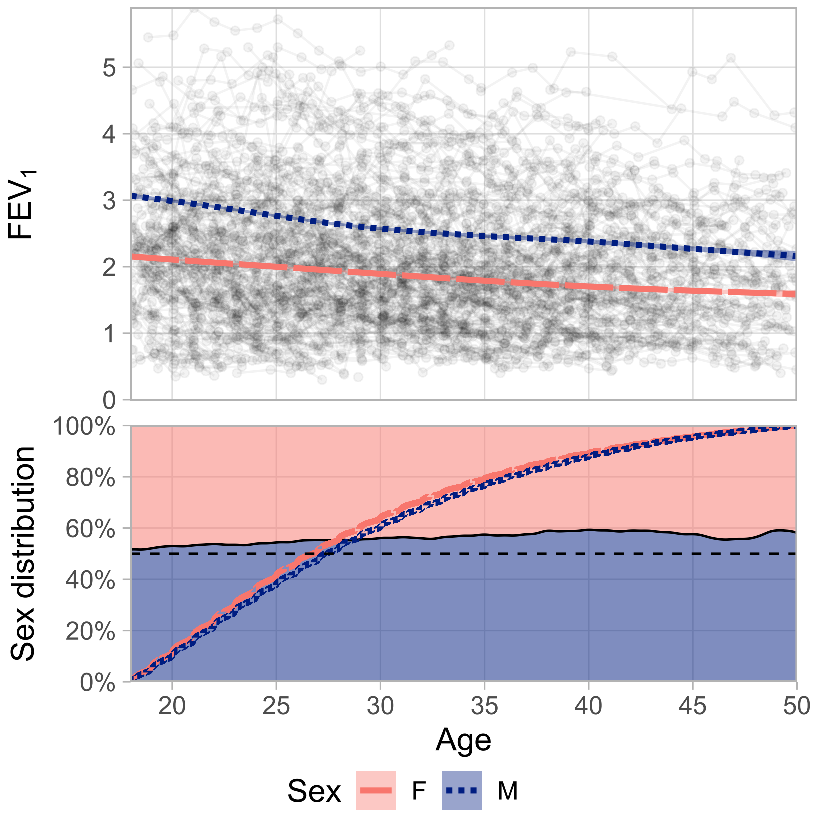


Figure 5: Top: Spaghetti plot for $FEV_1$ vs. age. Each black line connects the dots representing the lung function measurements for each individual at each annual review. The smoothed lines depicts the trend for females (red) and males (blue) with corresponding confidence intervals. For plotting clarity, only a random subset of dots and lines is reported.
Bottom: Sex distribution in the dataset selected for the analysis. The red and blue lines show the empirical cumulative distribution function over age for females (red) and males (blue), giving at each age level $a$ the percentage of individuals younger than $a$. The black solid line shows the relative percentage of males and females at each age point. The black dashed line marks the 50% level.

#### B.1 Results about interactions, predictions and random effects

The parametric term for sex (accounting for the average difference between male and female smooth functions) and the corresponding 95% credible interval is equal to 0.10 (0.08; 0.12) in the mean submodel and 0.25 (0.22; 0.27) in the variability submodel. These values can be observed as the average difference between the lines displayed in Figure 1.

Given these parameter estimates, we can predict the $FEV_1$ mean and standard deviation trajectories for specific covariate values. Some results are shown in Figure 6.

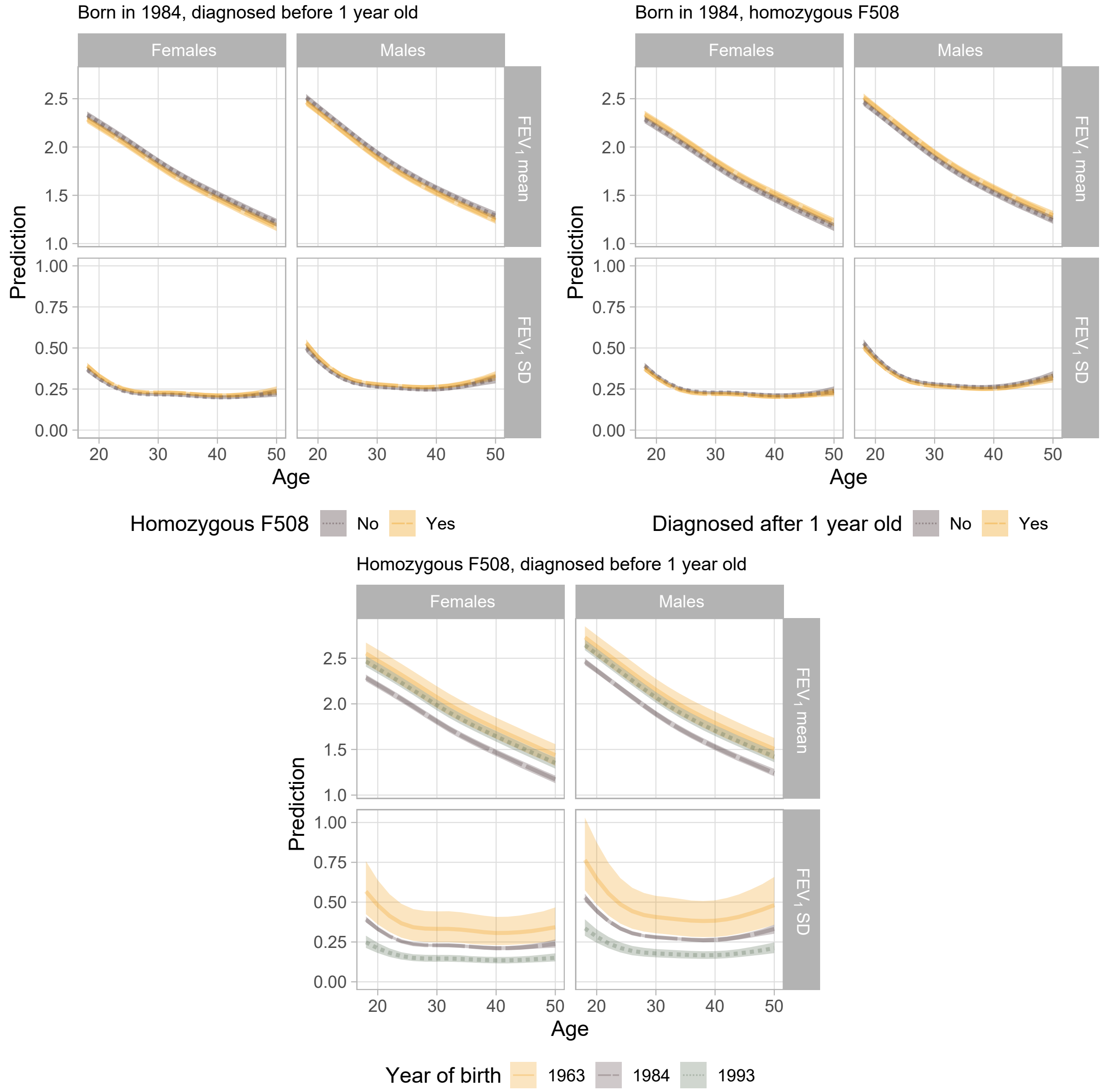


Figure 6: Posterior predictions of mean and standard deviation of $FEV_1$ by sex and age for specific sets of covariate values. Year 1984 is the median year of birth in the population.

The parameters of the distribution of the random effect matrix are also estimated (Table 2). For the mean submodel, the standard deviation of the random intercept is equal to 1.02. This means that if we consider a normal distribution with mean zero and this standard deviation, 95% of the individuals will get a predicted random effect in the range of (plus or minus) $2 \cdot 1.04 = 2.08$ $FEV_1$ litres. The posterior mean of the standard deviation of the scale random effect is equal to 0.45 and most of the posterior density lies far from the lower bound of 0 (see Figure 7 in Appendix B for the full posterior distribution), indicating that there is some heterogeneity that is not captured by the covariates in the model.

Looking at the individual estimates of the random effects jointly for location and scale (Figure 8), the point cloud is centered around 0 as per the model specification (statistical model described in Equation (3)). The slope of the least squares regression of the location random effects onto the scale random effects is positive and the sign agrees with the correlation parameter in Table 2.

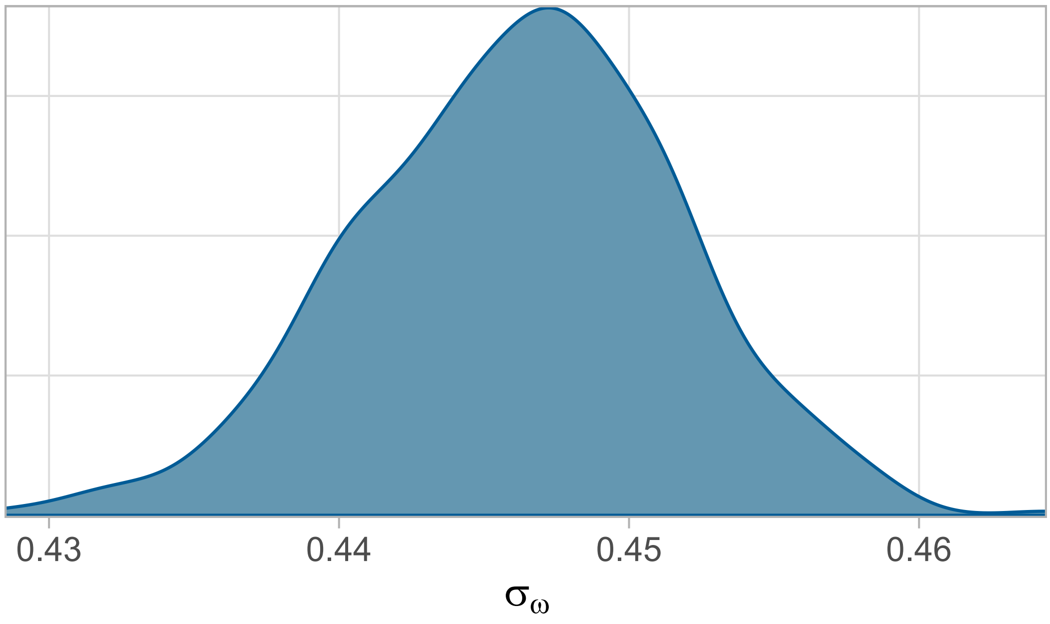


Figure 7: Posterior density of the standard deviation of the scale random effect $\sigma_\omega$.

The relationship between scale random effect estimates and sample size is displayed in Figure 9. A noteworthy result is obtained for individuals with low number of annual reviews, for which the scale random effects remain closer to zero. The model picks more information about the subject-specific deviation from common (fixed-effects) variability when the number of reviews increases.

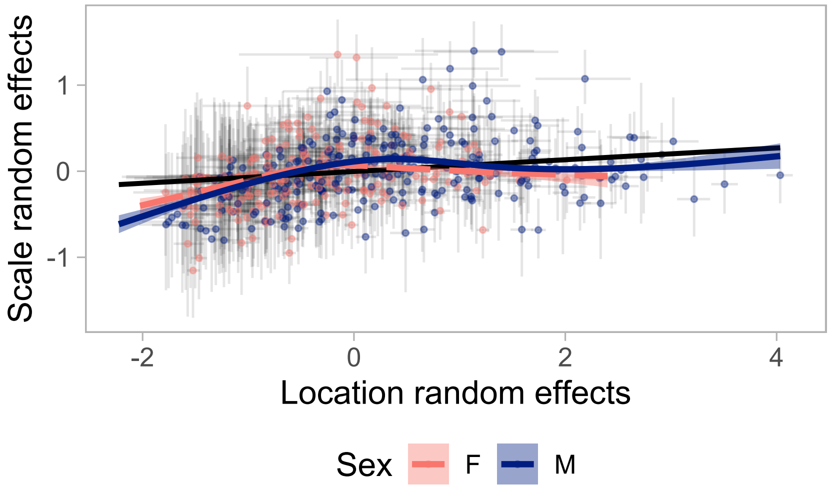


Figure 8: Plot of the relationship between location and scale random effects. Each individual is represented by a dot and a cross. The dot represents the point estimates for the individual location and scale random effects. The horizontal line of the cross refers to the 95% posterior interval for the location random effect, while the vertical line indicates the same interval for the scale random effects. The black line is the OLS fit for the regression of the location random effect onto the scale random effects, while the blue and red line give the nonparametric trend for males and females, respectively. For plotting clarity, only a subset of dots and crosses is reported.

#### B.2 Model diagnostics

| Covariate | $\hat{R}$ |
|---|---|
| **Mean submodel** | |
| Intercept | 1.01 |
| Sex main term (Male) | 1.02 |
| Diagnosis after 1 years old | 1.00 |
| Homozygous for F508 | 1.01 |
| **Variability submodel (log SD)** | |
| Intercept | 1.00 |
| Sex main term (Male) | 1.00 |
| Diagnosis after 1 years old | 1.00 |
| Homozygous for F508 | 1.00 |
| **Random effect distribution** | |
| SD for location random effects | 1.05 |
| SD for scale random effects | 1.00 |
| Correlation between random effects | 1.00 |

Table 3: Diagnostic checks on model estimates. $\hat{R}$ is the Gelman-Rubin ratio convergence diagnostic. When $\hat{R}$ is much higher than 1, it indicates that convergence has not been reached for that parameter and estimates are unreliable.

As a graphical assessment of quality of fit, the simulated posterior densities are plotted against the observed $FEV_1$ distribution (Figure 10). This procedure is an example of a posterior predictive check [18], that aims at evaluating in a qualitative way whether the model can generate data which resemble the observed data. The overall shape is mostly captured, although some discrepancy occurs in the lower part of the domain.

A model with an additional parameter for skewness (i.e. with a skew-normal distribution with the

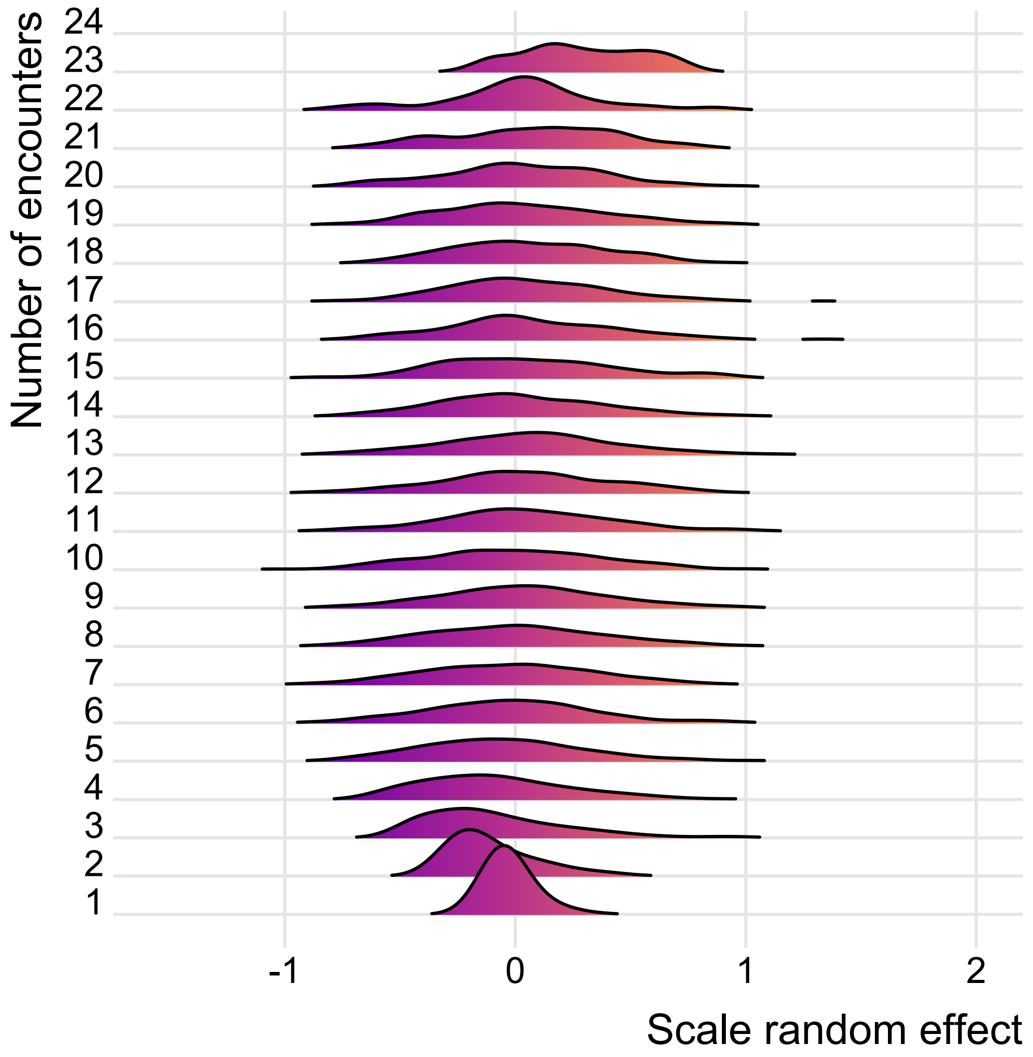


Figure 9: Distributions of scale random effects grouped by number of annual reviews per individual.

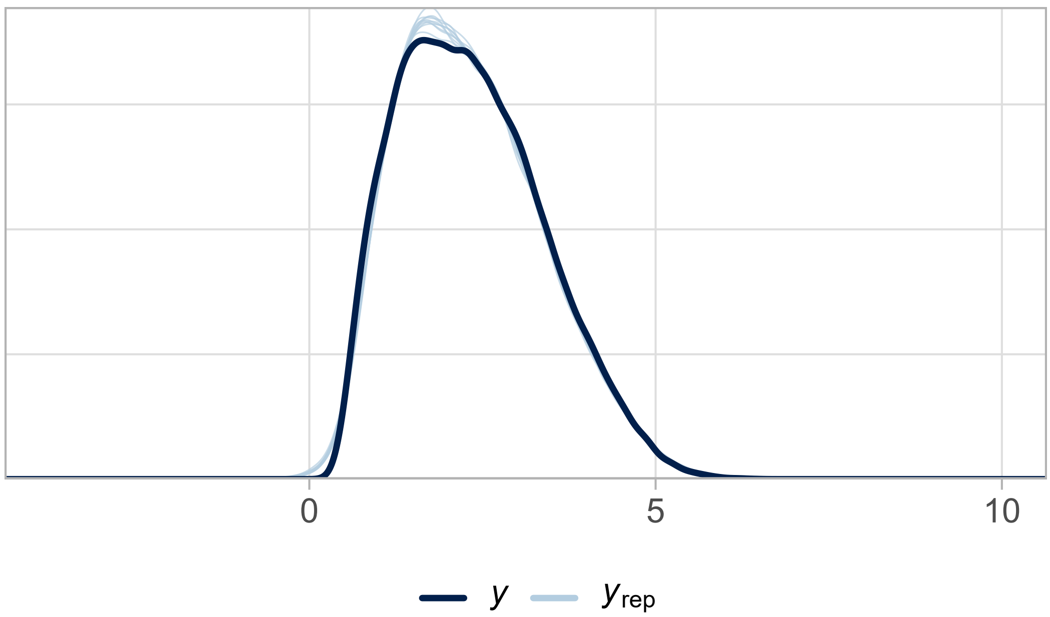


Figure 10: Posterior predictive checks. The dark blue line represents the density of the outcome ($FEV_1$) in the dataset, while the lighter lines are 10 simulated posterior distributions generated from the model.

same specification for mean and variance) did not improve the fit (results not reported here).
